# CD276 in Meningioma Transcriptomic Classification: Internal Development, External Validation, and Stability-Informed Interpretation

**DOI:** 10.64898/2026.04.03.26350116

**Authors:** Hyojae Lee, Hyeoneui Kim

**Affiliations:** College of Nursing, Seoul National University, Seoul, Republic of Korea

**Keywords:** Meningioma, CD276, Transcriptomic classification, External validation, Calibration, Robustness

## Abstract

**Background:** CD276 has been proposed as a candidate gene associated with the biological characteristics of meningioma, but its predictive position and interpretive significance within a transcriptomic classifier have not yet been clearly established. Accordingly, this study aimed to evaluate CD276 stepwise across internal model development, external validation, calibration, decision-analytic assessment, feature stability, and robustness analyses using public transcriptomic cohorts.

**Methods:** The analyses in this study were organized into two interconnected notebooks. In Notebook A, we reconstructed the internal training cohort (GSE183653), evaluated the CD276 single-gene signal, and then developed a transcriptome-wide multigene classifier. We also performed permutation importance, bootstrap confidence interval, label permutation test, repeated cross-validation, CD276 ablation, and internal calibration analyses. In Notebook B, we reproduced the external validation cohort (GSE136661) in a fixed common-gene space, applied train-only recalibration and train-only threshold transfer, and extended the interpretation through decision curve analysis, stability analysis, enrichment analysis, and one-factor-at-a-time robustness analysis.

**Results:** The internal training cohort consisted of 185 samples and 58,830 genes, of which 25 were WHO grade III cases. CD276 expression showed a significant association with WHO grade, but the internal discrimination of the CD276-only baseline was limited (ROC-AUC 0.628, average precision 0.323, balanced accuracy 0.540). In contrast, the initial transcriptome-wide model showed ROC-AUC 0.834 and PR-AUC 0.509, and under 5-fold cross-validation, the canonical full-transcriptome model and the CD276-forced 5,001-feature branch showed mean ROC-AUC/PR-AUC of 0.854/0.564 and 0.855/0.606, respectively, outperforming the CD276-only baseline at 0.644/0.391. CD276 was not included in the initial 5,000-feature filtered set and ranked 900th among 5,001 features even in the forcibly included 5,001-feature branch. In paired ablation analysis, the performance difference attributable to inclusion of CD276 was effectively close to zero (delta ROC-AUC 0.000062, delta PR-AUC 0.000056). Internal calibration analysis showed an overconfident probability pattern (Brier score 0.10501, intercept -1.421392, slope 0.413241). In external validation, the fixed multigene pipeline achieved ROC-AUC 0.928 and PR-AUC 0.335. Train-only recalibration improved calibration metrics while preserving discrimination, and decision curve analysis showed threshold-dependent but limited external utility. Stability analysis showed overlap between core-stable genes and high-impact genes, but CD276 was not supported as a dominant stable core feature and remained in the target-of-interest tier. In robustness analysis, some perturbations preserved the primary interpretation, whereas others revealed transform sensitivity or an alternative high-performing feature-space solution.

**Conclusions:** CD276 is a gene of interest associated with meningioma grade, but it was difficult to interpret it as a strong standalone predictor or a dominant stable classifier feature. In this study, the main basis of predictive performance lay not in CD276 alone but in a broader multigene transcriptomic structure, and probability output needed to be interpreted conservatively with calibration taken into account. These findings position CD276 not as a direct single-gene classifier but as a biology-motivated target-of-interest that should be interpreted within a broader transcriptomic program.

## Introduction

Meningioma is a tumor group that shows heterogeneity in terms of clinical course and risk of recurrence, and WHO grade is used as an important criterion for distinguishing these differences. However, because morphological classification alone is sometimes insufficient to fully explain the biological aggressiveness or prognosis of the tumor, interest has continued to grow in transcriptomic features and predictive models that can reflect molecular characteristics. In particular, analyses using public transcriptomic cohorts provide a useful foundation for developing reproducible classifiers and establishing external validation frameworks.

In this context, CD276 may be considered a gene of interest associated with the grade-associated biology of meningioma. Indeed, in the internal training cohort of this study, CD276 expression showed a significant association with WHO grade, and this relationship was maintained even after log1p transformation or winsorization. However, the fact that a specific gene is associated with tumor grade does not necessarily mean that it is a strong standalone predictor or a dominant feature within a multigene classifier. Predictive utility needs to be evaluated jointly in relation to single-gene signal, broader transcriptomic structure, probability calibration, threshold-dependent utility, and feature stability.

In the context of this study, two issues were particularly important. First, whether the discrimination observed in the internal cohort would also be maintained in the external cohort was a question of generalizability. Second, whether the fact that a feature was included in the model would actually translate into branch-level contribution or stable biological interpretation was another key question. In many cases, even when biologically interesting candidate genes exist, the main basis of predictive performance may be formed by a structural combination of multiple genes rather than by a single marker. In addition, even when discrimination is high, if the probability scale is not well calibrated, additional correction is required for threshold-based decision making or actual risk stratification. Therefore, it is important to adopt an approach that evaluates single-gene association, multigene discrimination, external validation, calibration, decision-analytic utility, stability, and robustness together rather than in isolation.

Based on these considerations, this study aimed to evaluate stepwise the predictive position and interpretive significance of CD276 using public GEO meningioma transcriptomic cohorts. To this end, the analyses were divided into two interconnected notebooks. In Notebook A, we reconstructed the internal training cohort and performed CD276 single-gene analysis, transcriptome-wide multigene classifier development, feature-space construction, permutation importance, bootstrap confidence interval, label permutation test, repeated cross-validation, CD276 ablation, and internal calibration. Subsequently, in Notebook B, we reproduced external validation in a fixed common-gene space, applied train-only recalibration and train-only threshold transfer, and extended the evaluation of predictive performance and interpretability through decision curve analysis, stability analysis, enrichment analysis, and one-factor-at-a-time robustness analysis. Through this design, the present study sought not only to view CD276 as a simple grade-associated marker, but also to examine in a more conservative and reproducible manner what role it plays within an actual transcriptomic classifier and how it should be understood in relation to a broader multigene program.

## Methods

The analyses in this study were divided into two independent notebooks according to their respective roles. Notebook A was responsible for the development and internal validation stage, including reconstruction of the internal training cohort, restoration of WHO grade labels, CD276 single-gene analysis, construction of a transcriptome-wide internal classifier, evaluation of feature importance, comparison of the CD276-forced branch, and bootstrap, permutation, repeated cross-validation, ablation, and internal calibration analyses. In contrast, Notebook B was responsible for the stage of external validation and extended interpretation, in which external cohort validation was reproduced in an aligned common-gene space based on the input structure and analytical rules established in Notebook A, followed by train-only recalibration, threshold selection, decision curve analysis, permutation-based external null analysis, stability analysis, enrichment analysis, robustness stress testing, and cross-axis integrative interpretation. Accordingly, Notebook A was designed to address internal model development and methodological validation, whereas Notebook B was designed to address external generalizability and the extension of biological and analytical interpretation.

## Methods-A

### Selection and Reconstruction of the Internal Training Cohort

Notebook A began by identifying, among public GEO meningioma transcriptomic cohorts, those for which sample-level WHO grade labels could be recovered and for which a reproducible expression-data pipeline could be constructed using only GEO-provided files. Prespecified candidate GEO series were evaluated for the recoverability of WHO grade labels and the availability of usable expression data, and GSE183653 was ultimately selected as the internal training cohort and GSE136661 as the external validation cohort. For GSE183653, series matrix metadata and TPM matrix sample keys were aligned to restore sample-level WHO grade labels, and the internal training cohort consisted of 58,830 genes and 185 samples, including 86 grade I, 74 grade II, and 25 grade III cases. Thereafter, WHO grade III status was defined as the binary outcome.

### CD276 Single-Gene Analysis

CD276 expression values were extracted from the aligned GSE183653 TPM matrix and merged with WHO grade metadata, after which the association between CD276 and WHO grade was evaluated. Differences across the three grade groups were assessed using the Kruskal–Wallis test, and pairwise differences between WHO grades were assessed using the Mann–Whitney U test. In addition, the same nonparametric tests were repeated after applying log1p transformation and top 1% winsorization in addition to the original TPM scale, in order to examine whether the observed association depended on the value scale. The CD276 single-gene baseline was defined as a logistic regression classifier using CD276 expression alone as input and WHO grade III status as the outcome, and was evaluated by 5-fold stratified out-of-fold prediction with ROC-AUC, PR-AUC, confusion matrix, and balanced accuracy.

### Transcriptome-Wide Internal Classifier and Feature-Space Construction

For multigene internal classifier analysis, the full GSE183653 TPM expression matrix was realigned to construct a transcriptome-wide input matrix. Log1p transformation was applied to the full input matrix, and a reduced transcriptome matrix was generated by selecting the top 5,000 genes according to gene-wise variance. The initial transcriptome-wide classifier was defined as a pipeline using this 5,000-feature matrix as input and consisting of StandardScaler(with_mean=True, with_std=True) and elastic-net penalty logistic regression (solver=“saga”, l1_ratio=0.5, C=1.0, max_iter=5000, class_weight=“balanced”, random_state=42). Evaluation was performed using 5-fold stratified out-of-fold prediction, and ROC-AUC, PR-AUC, accuracy, balanced accuracy, precision, recall, and F1 score were calculated from the OOF probabilities. Subsequently, a threshold grid spanning 0.05–0.95 was explored to characterize the internal operating-threshold landscape, and first-pass permutation importance was performed using the reduced matrix as input to assess the relative importance of CD276.

### Comparison of the CD276-Forced Branch and Internal Branches

To examine the possibility that CD276 might be excluded from the initial 5,000-feature filtered matrix, a separate multigene branch consisting of 5,001 genes was constructed by forcibly including CD276 in addition to the top 5,000 genes ranked by variance. Permutation importance was recalculated for this branch as well to assess the relative position of CD276. Three internal classifier structures were then compared. The first was a canonical full-transcriptome multigene baseline using all 58,830 transcriptomic features, the second was the 5,001-feature multigene branch, and the third was the CD276-only baseline. All three branches used WHO grade III status as the common outcome, and internal discrimination was compared according to differences in input feature space and feature composition while maintaining the class-balanced logistic regression family and 5-fold stratified cross-validation.

### Bootstrap, Permutation Null, and Repeated Cross-Validation

The internal performance uncertainty of the three branches was quantified using prediction-level bootstrap confidence intervals. For the canonical full-transcriptome multigene baseline, the 5001-feature multigene branch, and the CD276-only baseline, B=2000, SAVE_EVERY=5, BOOTSTRAP_RANDOM_SEED=42, and THRESHOLD_REF=0.5 were used. Branch-specific 5-fold OOF predicted probabilities were first generated, after which patient-level resampling based on the sample-level prediction state was repeatedly performed to estimate percentile-based 95% confidence intervals for ROC-AUC, PR-AUC, balanced accuracy, and accuracy. In addition, label permutation tests with B=500 were performed for the canonical full-transcriptome model and the 5001-feature multigene branch, respectively, to evaluate whether the observed performance exceeded a random label structure. Furthermore, repeated stratified cross-validation using C_GRID = [0.1, 0.3, 1.0, 3.0, 10.0] was performed separately for the two multigene branches to assess split-to-split variability and sensitivity to regularization strength. The canonical full-transcriptome model was evaluated using RepeatedStratifiedKFold(n_splits=5, n_repeats=20, random_state=42) and StandardScaler(with_mean=False), whereas the 5001-feature branch was evaluated under the same repeated stratified validation structure but with an elastic-net logistic regression including StandardScaler(with_mean=True, with_std=True).

### CD276 Ablation, Regularization-Strength Analysis, and Internal Calibration

To evaluate the contribution of CD276 to the performance of the 5001-feature multigene branch, paired ablation repeated cross-validation was performed by comparing the CD276-including branch and the CD276-excluding branch on the same splits using X2 as the basis. X_excl was defined as X_incl with only the CD276 column removed, and the same elastic-net logistic regression including C_GRID = [0.1, 0.3, 1.0, 3.0, 10.0], RepeatedStratifiedKFold(n_splits=5, n_repeats=20, random_state=42), and StandardScaler(with_mean=True, with_std=True) was applied identically to both branches. In addition, to evaluate how the CD276 coefficient and branch-level discrimination changed according to regularization strength within the CD276-forced 5001-feature branch, a joint regularization-strength analysis based on RepeatedStratifiedKFold(n_splits=5, n_repeats=10, random_state=42) was performed, and the CD276 coefficient was tracked together with branch-level ROC-AUC and PR-AUC in each split. Finally, internal calibration was evaluated using the OOF predicted probabilities of the initial transcriptome-wide multigene model. For this purpose, the Brier score was calculated, and calibration intercept and slope were estimated using a logistic calibration model with the logit-transformed probability values as a single predictor, followed by decile-based calibration analysis using 10 quantile bins.

## Methods-b

### External Validation Framework

The main evaluation stage of Notebook B was conducted by restoring the previously validated legacy analysis snapshot and then reproducing external validation in an aligned common-gene space. The restored state included pre-aligned expression matrices for the training cohort and the external validation cohort, binary outcome labels, the common gene axis, and previously generated external-prediction-related objects. The external validation cohort consisted of a total of 160 samples, of which 7 were positive cases. All external evaluations were performed in a fixed common-gene space shared with the training cohort.

The final classifier was implemented as an L1-penalized logistic regression using the saga solver. The modeling pipeline included zero-variance feature removal and standardization before classifier fitting. The regularization strength was fixed at C=1.0, and the maximum number of iterations was set to 8000. Performance in the external cohort was summarized using ROC-AUC, PR-AUC, accuracy, balanced accuracy, and threshold-dependent operating characteristics.

### Calibration and Threshold Selection

To avoid bias arising from threshold optimization based on external data, calibration and threshold selection were performed using the training data only. Out-of-fold predicted probabilities were generated in the training cohort using 5-fold 5-repeat repeated stratified cross-validation, and a logistic recalibration model was fitted using the logit-transformed probability values as input. This recalibration model was then applied to the predicted probabilities in the external cohort.

Operating thresholds were also selected using only the out-of-fold calibrated probabilities from the training cohort. The selection rules included the threshold that maximized the F1 score, the threshold that maximized Youden’s J, the threshold that maximized specificity while satisfying sensitivity ≥ 0.80, and the reference threshold of 0.50. These thresholds derived from the training data were then applied to the externally calibrated probabilities to obtain clinically interpretable operating points.

### Decision Curve Analysis

Clinical utility in the external cohort was evaluated using decision curve analysis based on external predicted probabilities after train-only recalibration. Net benefit was calculated over the prespecified threshold range of 0.01–0.50 and compared with the treat-all and treat-none strategies. Uncertainty was quantified by patient-level bootstrap resampling (B=2000, seed 42), and 95% confidence intervals for threshold-specific net benefit were estimated. Net benefit and interventions avoided were also converted to a per-100-patients basis to provide a clinical-impact summary. To assess whether the decision structure identified during model development was maintained in the external cohort, Train (OOF) and External decision curves were overlaid and compared.

### Permutation-Based Null Analysis

To evaluate whether the observed external validation performance exceeded the level expected under a random label structure, a train-label permutation null analysis evaluated on the external cohort was performed. The training labels were first permuted, after which the same classification pipeline was fitted and ROC-AUC and PR-AUC were calculated in the external cohort to construct a null distribution. This analysis was then extended by increasing the number of permutations to estimate the empirical null distribution more stably. The observed performance was compared with the permutation null distribution to calculate empirical permutation p-values, thereby assessing whether the external validation performance exceeded the level expected under a random label structure.

### Stability Analysis

Feature stability was evaluated through bootstrap-based stability analysis in the validated common-gene space. Two independent seed runs (42 and 2025) were performed, and each run consisted of 500 bootstrap replicates. In each replicate, class-stratified resampling was performed on the training cohort, after which the L1-penalized logistic regression model was refitted and performance was evaluated in the external cohort. During the iterative process, selection frequency and coefficient-based summary statistics were accumulated for each gene, and for the gene of interest, CD276, a separate coefficient trace was tracked. All stability analyses were performed on the same validated common-gene axis in order to preserve comparability across seeds and across downstream interpretations.

At the interpretation stage, the two stability runs were integrated to summarize features. Core stable genes were conservatively defined as genes that showed high repeated selection frequency in both seed runs and concordant mean coefficient direction. High-impact genes were ranked using a coefficient magnitude-based importance metric, and CD276 was separately tracked as a prespecified target-of-interest gene regardless of whether it met the core stability criterion.

### Enrichment Analysis

Biological interpretation of the stability analysis results was performed on the basis of prespecified enrichment analyses. The background gene universe was fixed as the entire validated common-gene axis (31,582 genes). Over-representation analysis was performed primarily for core stable genes and secondarily for the overlap set between core stable genes and high-impact genes. In addition, ranked enrichment analysis based on a stability-informed ranking across all genes was performed. In this ranked analysis, a stability-weighted score defined as the product of mean selection frequency and mean absolute coefficient magnitude was used. The enrichment databases used were GO Biological Process, Reactome, and Hallmark. Multiple-testing correction was performed using the Benjamini–Hochberg false discovery rate, and the significance threshold was defined as FDR < 0.05.

Because the stability checkpoints did not store per-bootstrap sign count information, exact sign-frequency-based directional enrichment analysis was not performed. Accordingly, directionality was interpreted conservatively only and was not used to define sign-separated enrichment gene sets.

### Robustness Analysis

Robustness evaluation was conducted within a one-factor-at-a-time framework in which only a single component of the primary pipeline was varied at a time. Five robustness axes were considered, reflecting the duplicate-symbol aggregation rule (Axis A), external input transformation rule (Axis B), common-gene quality-control restriction (Axis C), convergence setting (Axis D), and feature prefilter rule (Axis E), respectively. Each axis was run with two independent seeds (42 and 2025), and all robustness runs used 500 bootstrap replicates.

Robustness results were not pooled into the primary stable gene definition or the primary biological interpretation. Instead, these analyses were used as stress tests to evaluate whether the primary predictive and biological themes were maintained under analytic perturbation. The main reporting targets were predictive drift in the external cohort, overlap with the primary core gene set, recurrence of pathway-level themes, and changes in the relative position of CD276.

### Cross-Axis Integrative Interpretation

After generating axis-specific robustness summaries, all robustness axes were integrated to compare gene-level concordance, pathway-level consistency, and the behavior of CD276 with the primary interpretation. Concordance was summarized using overlap fractions and Jaccard indices between axis-specific core gene sets and overlap sets. CD276 was evaluated in all robustness runs on the basis of a selection-frequency proxy and a coefficient-magnitude proxy.

This integrative analysis showed that robustness results should be interpreted not as a single homogeneous summary, but rather in terms of how the primary interpretation was maintained or altered under different analytic perturbations. Accordingly, robustness analysis was interpreted not as an alternative primary analysis, but as a supplementary validation framework for assessing the context and defensibility of the primary findings.

## Results-A

### Reconstruction of the Internal Training Cohort and the CD276 Single-Gene Signal

Reconstruction of the internal training cohort selected as GSE183653 showed that the final analytic set consisted of a total of 185 samples and 58,830 genes, with a WHO grade distribution of 86 grade I, 74 grade II, and 25 grade III cases. Thereafter, the binary outcome for internal analyses was defined as WHO grade III status.

Upon first examining the CD276 single-gene signal, the same grade distribution was retained in the sample-level training table, and the range of CD276 expression values was 1.67–265.16 TPM. The overall difference in CD276 expression across WHO grades was significant by the Kruskal–Wallis test (statistic=7.122, p=0.028), and the mean and median values in the grade III group were 69.38 TPM and 52.28 TPM, respectively, both higher than those in the grade I group (41.51 TPM and 36.65 TPM) and the grade II group (46.82 TPM and 43.13 TPM). In pairwise comparisons, the difference between grade I and grade III remained significant after correction (Bonferroni-corrected p=0.034), whereas the difference between grade II and grade III showed a difference at the level of the raw p-value but was not significant after correction (raw p=0.046, corrected p=0.139). (Figure 1)

**Figure 1.**
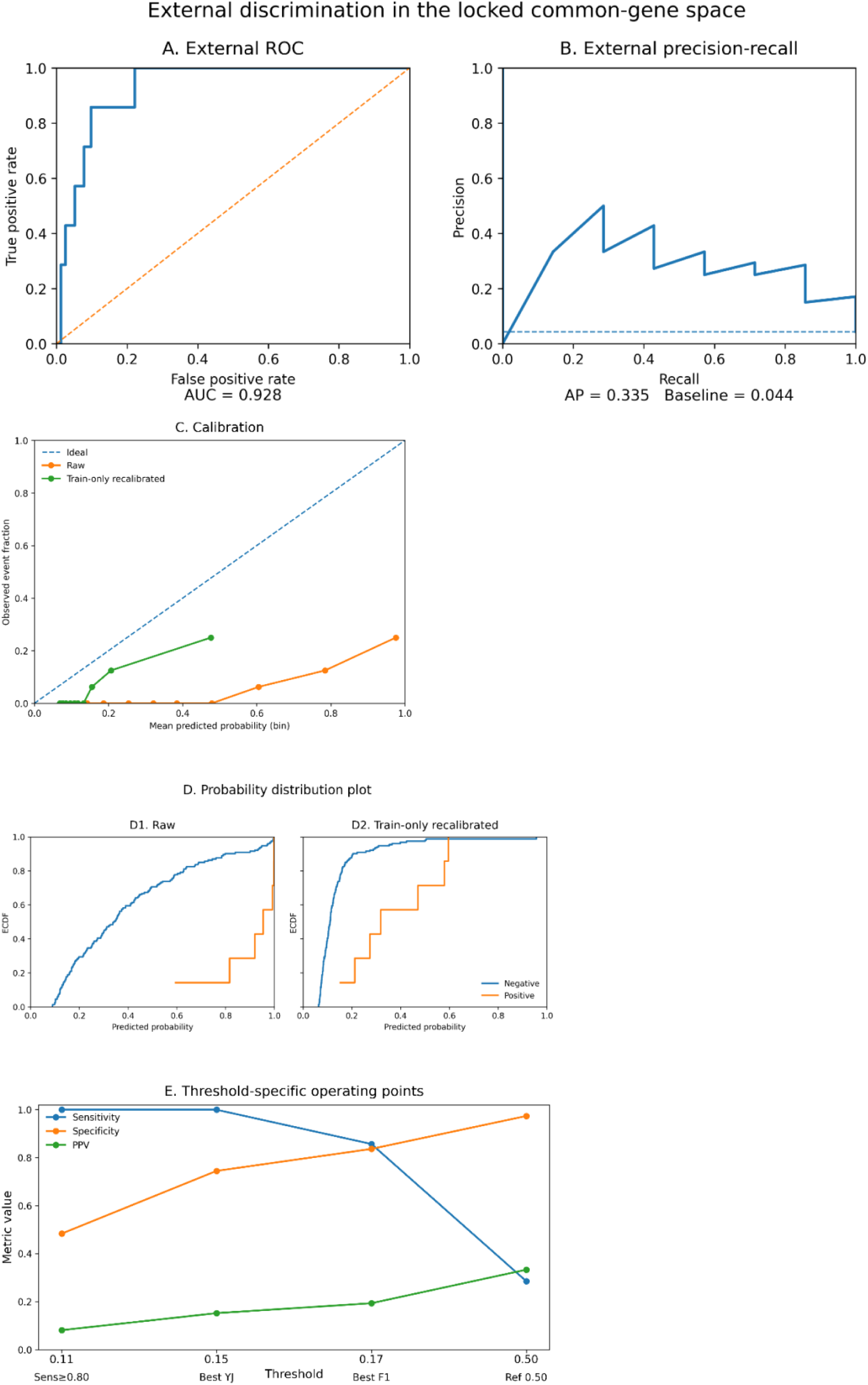
Summary of external validation, calibration, and operating thresholds. (A) ROC curve of the external cohort evaluated in a fixed common-gene space. (B) Precision-recall curve for the same external predictions, with external event prevalence indicated as the baseline. (C) Comparison of calibration curves for raw external probabilities and train-only recalibrated probabilities; the diagonal line indicates ideal calibration. (D) Predicted probability distributions of external negative and positive samples before and after recalibration, presented in the form of empirical cumulative distributions. (E) Summary of sensitivity, specificity, and positive predictive value for thresholds 0.11, 0.15, and 0.17 selected by train-only rules, together with the reference threshold of 0.50. The recalibration model was fitted using only the repeated out-of-fold predictions of the training cohort, and external data were not used for tuning.

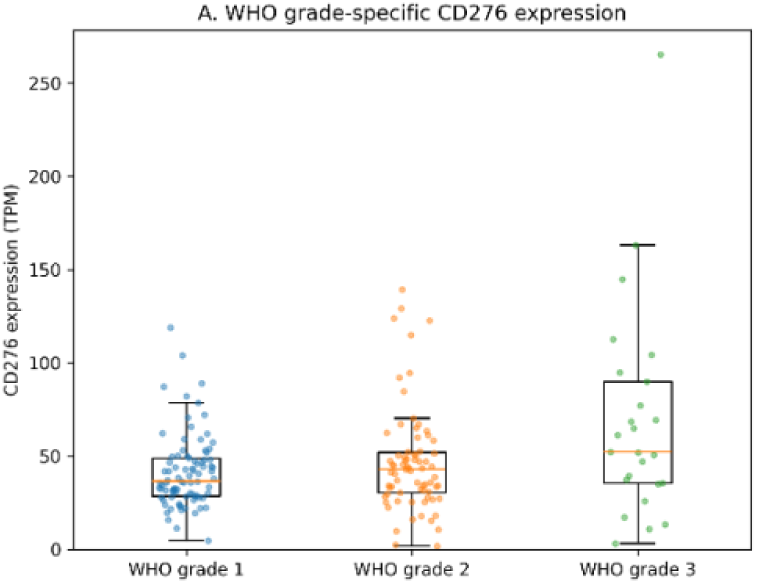

This directional pattern was maintained even after the prespecified value transformations. After log1p transformation and top 1% winsorization, the Kruskal–Wallis statistic and p-value were essentially unchanged (statistic≈7.122, p=0.028), and the median value in the grade III group remained consistently higher than those in grade I and grade II. For example, on the log1p scale, the median value in the grade III group was 3.98, whereas those in grade I and grade II were 3.63 and 3.79, respectively. After winsorization, the median value in the grade III group also remained 52.28.

However, the internal discrimination of the 5-fold out-of-fold single-gene baseline classifier using CD276 alone was limited. This model showed ROC-AUC 0.628, average precision 0.323, and balanced accuracy 0.540, and at the reference threshold of 0.5, the confusion matrix was TN 160, FP 0, FN 23, and TP 2. In other words, only 2 of the 25 grade III cases were recovered, indicating that CD276 as a single feature did not provide sufficient classification performance in the internal training cohort.

### Internal discrimination of the transcriptome-wide multigene model

The analytic focus was then expanded to the whole-transcriptome level. After reconstructing the GSE183653 expression matrix as a sample-aligned transcriptome input, log1p transformation and variance filtering were applied, and the initial transcriptome-wide model input was reduced to 5,000 features. The 5-fold out-of-fold performance of this initial elastic-net logistic-regression model was ROC-AUC 0.834, PR-AUC 0.509, accuracy 0.859, and balanced accuracy 0.750. At the reference threshold of 0.5, the confusion matrix was TN 144, FP 16, FN 10, and TP 15, indicating higher internal discrimination than the CD276 single-gene baseline. (Table 1)

**Table 1.**
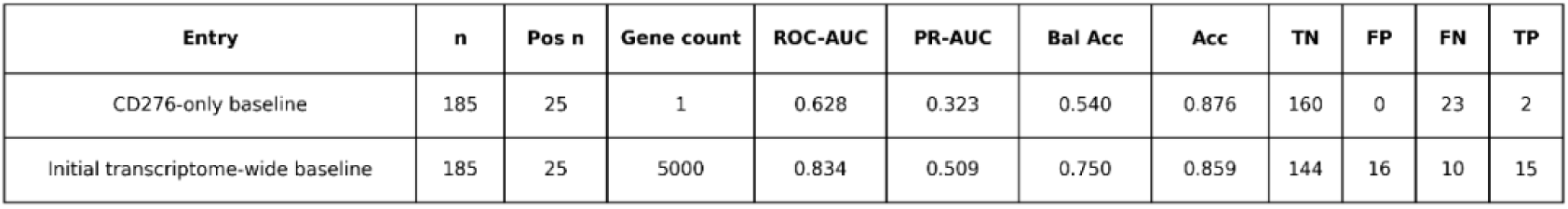
Internal baseline comparison.

| Entry | n | Pos n | Gene count | ROC-AUC | PR-AUC | Bal Acc | Acc | TN | FP | FN | TP |
| --- | --- | --- | --- | --- | --- | --- | --- | --- | --- | --- | --- |
| CD276-only baseline | 185 | 25 | 1 | 0.628 | 0.323 | 0.540 | 0.876 | 160 | 0 | 23 | 2 |
| Initial transcriptome-wide baseline | 185 | 25 | 5000 | 0.834 | 0.509 | 0.750 | 0.859 | 144 | 16 | 10 | 15 |

**Table 1.** Summary of external validation and recalibration External discrimination and calibration performance in the fixed common-gene space are summarized. Brier score, ECE10, recalibration intercept, and recalibration slope were compared between the raw probabilities and the train-only recalibrated probabilities, while ROC-AUC and PR-AUC remained unchanged after monotonic recalibration.

| Cohort | n | Pos n | ROC-AUC | PR-AUC | Brier (raw) | Brier (recal) | ECE10 (raw) | ECE10 (recal) | Cal intercept (raw) | Cal slope (raw) | Cal intercept (recal) | Cal slope (recal) |
| --- | --- | --- | --- | --- | --- | --- | --- | --- | --- | --- | --- | --- |
| External | 160 | 7 | 0.928 | 0.335 | 0.221 | 0.052 | 0.380 | 0.110 | -3.378 | 0.300 | -1.848 | 0.359 |

Upon examining permutation importance in the initial reduced feature space, CD276 was not included in the top 5,000 filtered gene set. Accordingly, a separate 5,001-feature multigene branch was constructed in which CD276 was forcibly included. When permutation importance was re-evaluated in this branch, CD276 ranked 900th among the 5,001 features, and both its importance mean and standard deviation were 0. In contrast, the top-ranked features in the same analysis included LTF, NCALD, MRFAP1, C4B, and MRFAP1L1.

When the three internal classifier structures were directly compared, both the canonical full-transcriptome multigene baseline and the 5,001-feature multigene branch maintained superior internal discrimination relative to the CD276-only baseline. Under 5-fold cross-validation, the mean ROC-AUC and PR-AUC of the canonical full-transcriptome baseline were 0.854 and 0.564, respectively, whereas those of the 5,001-feature multigene branch were 0.855 and 0.606. In contrast, the corresponding mean ROC-AUC and PR-AUC of the CD276-only baseline were 0.644 and 0.391. In addition, the mean balanced accuracy and mean accuracy were 0.641 and 0.876 for the canonical full-transcriptome model, 0.695 and 0.881 for the 5,001-feature branch, and 0.648 and 0.741 for the CD276-only baseline. Thus, in the internal training cohort, multigene transcriptomic structure provided substantially stronger discrimination than the single-gene CD276, and the 5,001-feature branch preserved discrimination close to that of the canonical full-transcriptome baseline while providing a reduced feature space. (Figure 2)

**Figure 2.**
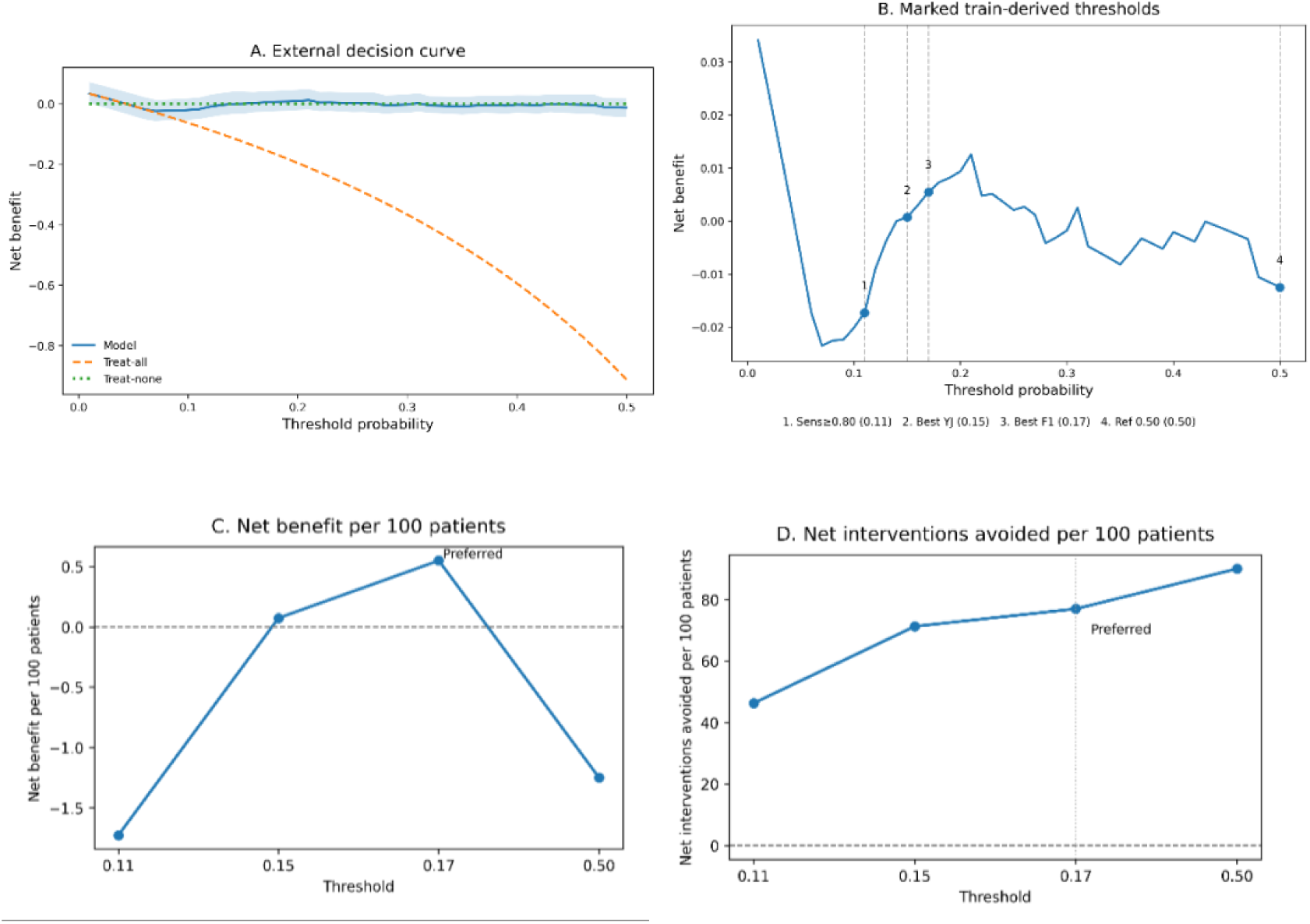
External decision-curve analysis. (A) External decision curve over the threshold probability range of 0.01–0.50, showing the model, treat-all, and treat-none strategies together with bootstrap confidence intervals. (B) Thresholds 0.11, 0.15, 0.17, and 0.50 selected by the train-only procedure are displayed on the external decision curve. (C) Net benefit per 100 patients at the displayed thresholds. (D) Net interventions avoided per 100 patients relative to treat-all at the same thresholds. Among the prespecified train-only candidate thresholds, only 0.17 showed the most favorable decision-analytic profile in the external cohort.

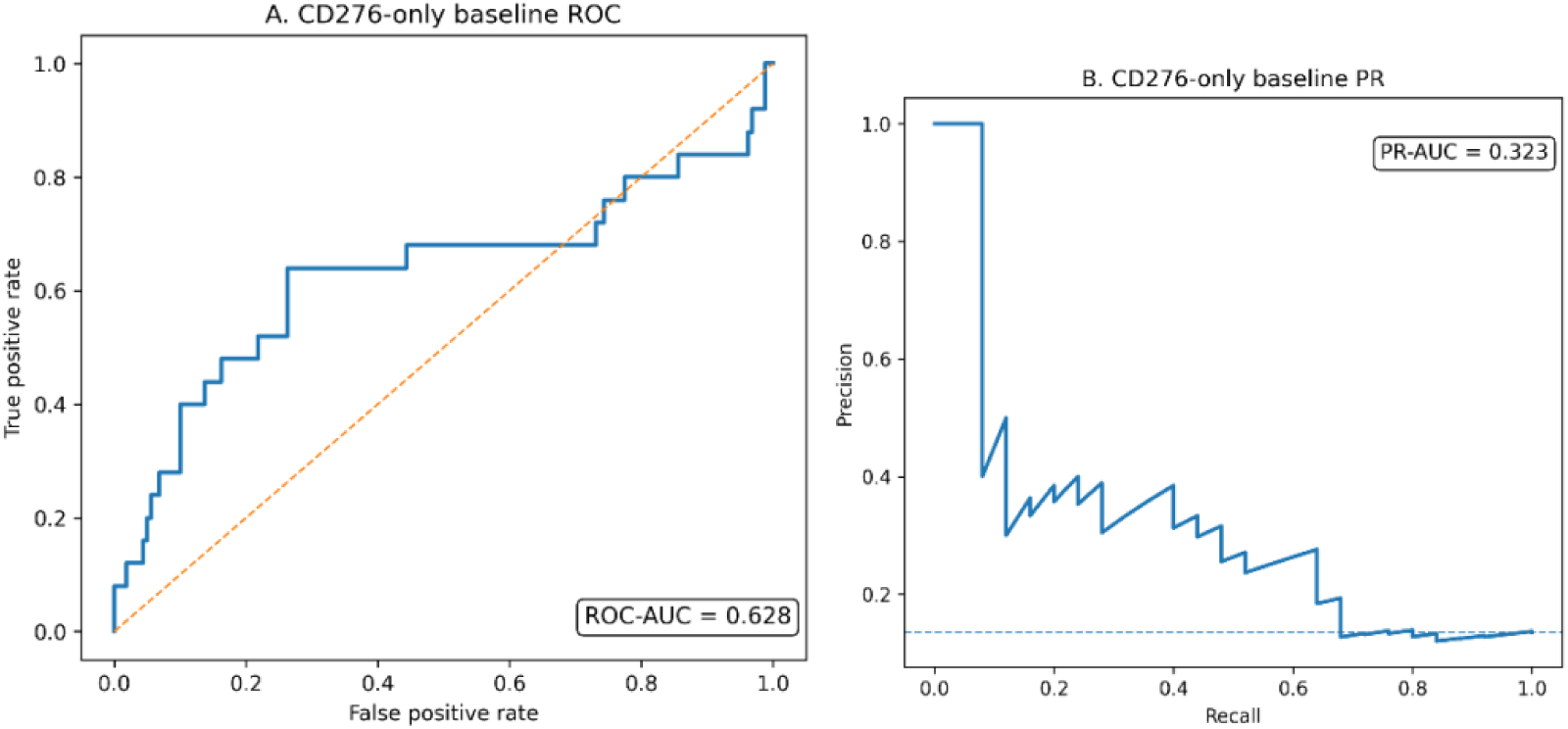

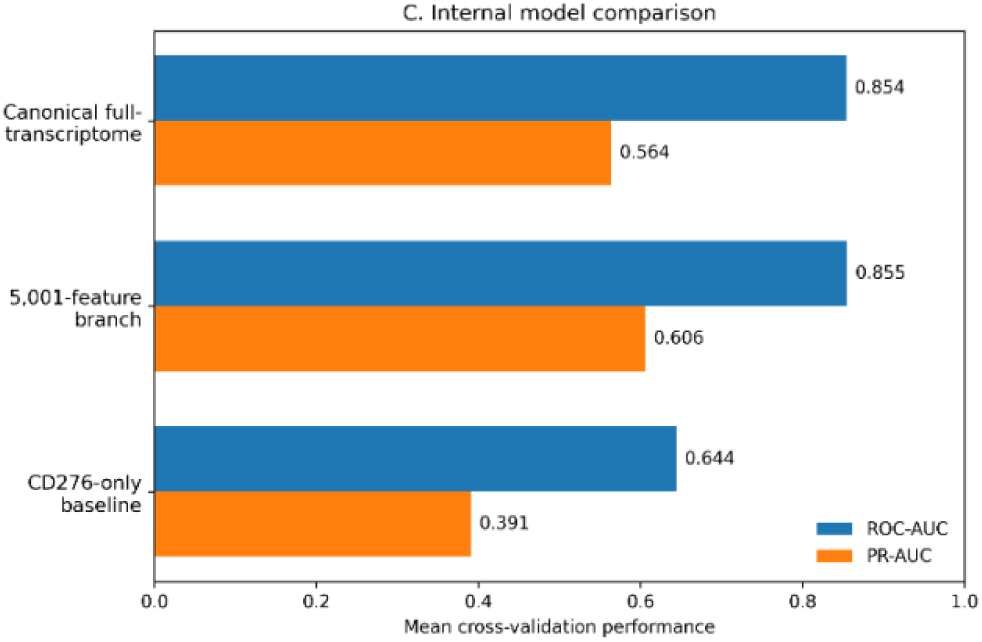

### Internal uncertainty, permutation null, and regularization sensitivity

When internal performance uncertainty was quantified by prediction-level bootstrap, the relative ordering among the three branches was maintained. The bootstrap summary for the canonical full-transcriptome multigene baseline was ROC-AUC 0.847 (95% CI, 0.753–0.926), PR-AUC 0.469 (0.311–0.691), balanced accuracy 0.641 (0.552–0.735), and accuracy 0.876 (0.827–0.919). The 5,001-feature multigene branch showed ROC-AUC 0.831 (0.725–0.920), PR-AUC 0.506 (0.335–0.726), balanced accuracy 0.695 (0.595–0.800), and accuracy 0.881 (0.832–0.924). In contrast, the CD276-only baseline showed ROC-AUC 0.626 (0.472–0.772), PR-AUC 0.319 (0.176–0.496), balanced accuracy 0.647 (0.547–0.753), and accuracy 0.741 (0.676–0.800). Thus, the superiority of the multigene models was maintained even after accounting for bootstrap uncertainty, and the 5,001-feature branch in particular showed a relatively favorable profile in terms of PR-AUC.

To examine whether the observed multigene performance could also be explained by a random label structure, label permutation tests were performed separately for the canonical full-transcriptome model and the 5,001-feature multigene branch. For both models, permutation p-values for ROC-AUC and PR-AUC were 0.001996, indicating that the internal multigene discrimination is difficult to explain solely by chance label alignment.

In the regularization-strength scan as well, the two multigene branches maintained generally stable performance ranges. In repeated stratified cross-validation of the canonical full-transcriptome model, the optimal C based on PR-AUC was 1.0, at which the mean ROC-AUC and PR-AUC were 0.837 and 0.578, respectively. By the same criterion, the optimal C for the 5,001-feature branch was 3.0, at which the mean ROC-AUC and PR-AUC were 0.819 and 0.576, respectively. The canonical model was slightly higher in ROC-AUC, whereas the 5,001-feature branch maintained nearly identical PR-AUC. In fact, across the full C-grid results of the canonical model, the mean ROC-AUC ranged from 0.815 to 0.844 and the mean PR-AUC ranged from 0.558 to 0.578, while in the 5,001-feature branch, the mean ROC-AUC ranged from 0.813 to 0.829 and the mean PR-AUC ranged from 0.552 to 0.576. (Table 2)

**Table 2.** Internal model summary.

| Model | Bootstrap<br>ROC-AUC<br>(95% CI) | Bootstrap<br>PR-AUC<br>(95% CI) | Bootstrap<br>Bal Acc<br>(95% CI) | Bootstrap<br>Acc<br>(95% CI) | Perm p<br>ROC-AUC | Perm p<br>PR-AUC | Selected<br>C | Repeated-CV<br>ROC-AUC | Repeated-CV<br>PR-AUC |
| --- | --- | --- | --- | --- | --- | --- | --- | --- | --- |
| Canonical full-transcriptome | 0.847 (0.753-0.926) | 0.469 (0.311-0.691) | 0.641 (0.552-0.735) | 0.876 (0.827-0.919) | 0.001996 | 0.001996 | 1.0 | 0.837 | 0.578 |
| 5,001-feature branch | 0.831 (0.725-0.920) | 0.506 (0.335-0.726) | 0.695 (0.595-0.800) | 0.881 (0.832-0.924) | 0.001996 | 0.001996 | 3.0 | 0.819 | 0.576 |
| CD276-only baseline | 0.626 (0.472-0.772) | 0.319 (0.176-0.496) | 0.647 (0.547-0.753) | 0.741 (0.676-0.800) | NA | NA | NA | NA | NA |

**Table 2.**
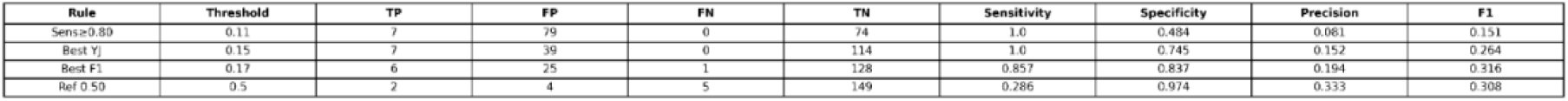
Threshold-specific operating points The confusion-matrix counts and operating metrics obtained when prespecified thresholds selected from the train-only calibrated training out-of-fold predictions were applied once to the externally recalibrated probabilities are summarized. TP, FP, FN, TN, sensitivity, specificity, precision, and F1 are presented for the sensitivity-oriented threshold (0.11), the best Youden’s J threshold (0.15), the best F1 threshold (0.17), and the reference threshold of 0.50.

### The branch-level contribution of CD276 was limited

To directly evaluate whether CD276 actually contributed to the performance of the 5,001-feature branch, paired ablation repeated cross-validation was performed by comparing the CD276-including branch and the CD276-excluding branch on the same splits. The most favorable row was observed at C=3.0, but the include–exclude difference was effectively close to zero. In this row, the mean ROC-AUC values of the include branch and the exclude branch were 0.791406 and 0.791344, respectively, and the mean PR-AUC values were 0.510400 and 0.510345, respectively. The mean delta ROC-AUC was 0.000062, and the mean delta PR-AUC was 0.000056, while the differences in balanced accuracy and accuracy were also zero. At other C values as well, the deltas remained at zero or at comparably negligible levels.

These results show that including CD276 in the feature space did not meaningfully increase branch-level discrimination. Therefore, CD276 was closer to a target that should be interpreted within a broader multigene transcriptomic context rather than a dominant feature determining the performance of the internal multigene classifier.

### Internal calibration assessment

Finally, internal calibration was evaluated for the OOF predicted probabilities of the initial transcriptome-wide multigene model. The Brier score of this model was 0.10501, the calibration intercept was -1.421392, and the calibration slope was 0.413241. This indicates that although the internal OOF predictions achieved a certain level of discrimination, the probability scale itself deviated from ideal calibration, with a slope substantially smaller than 1, reflecting an overconfident pattern.

This tendency was also confirmed in the decile-based calibration table. In the highest-risk bin, the mean predicted probability was 0.935341, whereas the observed event rate was only 0.578947, and the predicted probability range within the same bin was 0.793486–0.999998. In contrast, in the lower bins, the mean predicted probabilities were very low, ranging from 0.008 to 0.080, and the observed event rates also remained mostly at 0 or in the range of 0.053–0.111. Thus, the internal stage of Notebook A showed that the transcriptomic multigene model established meaningful discrimination while also indicating that the probability output should be interpreted conservatively from a calibration perspective. (Figure 3; Table 3)

**Figure 3.**
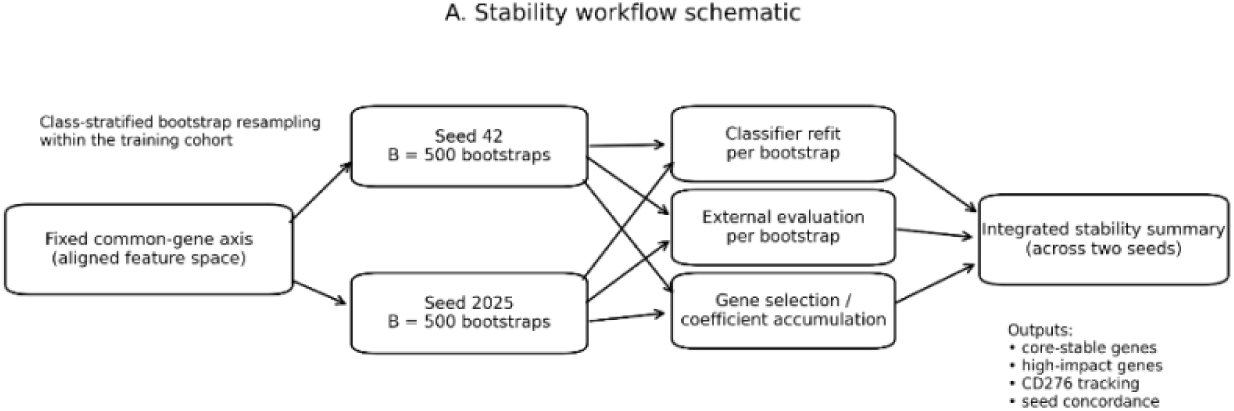
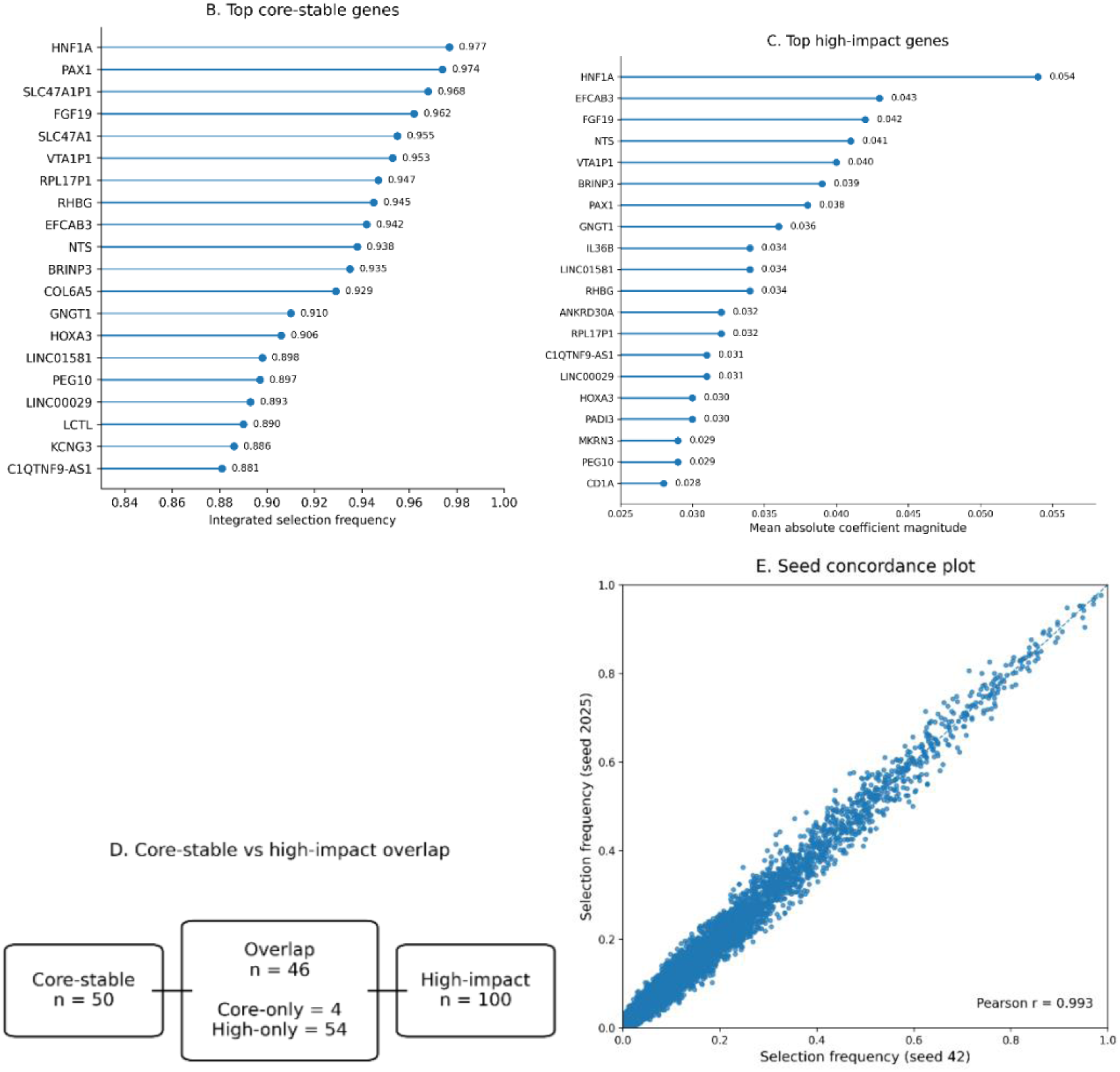
Stability-informed transcriptomic interpretation. (A) Stability workflow schematic using two completed seed schedules. (B) Top core-stable genes ranked by integrated selection frequency. (C) Top high-impact genes ranked by mean absolute coefficient magnitude. (D) Schematic overlap between the core-stable set and the high-impact set. (E) Gene-level selection frequency concordance plot between the two seed schedules. These results show a substantial overlap between repeated selection stability and effect magnitude and suggest that CD276 was not supported as a dominant stable core feature but remained in the target-of-interest tier.

**Table 3.**
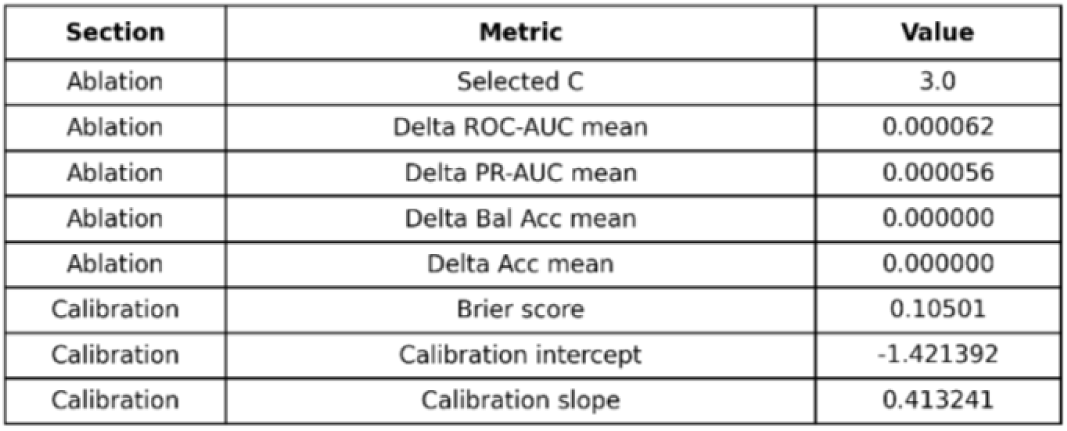
Ablation and calibration summary.

**Table 3.**
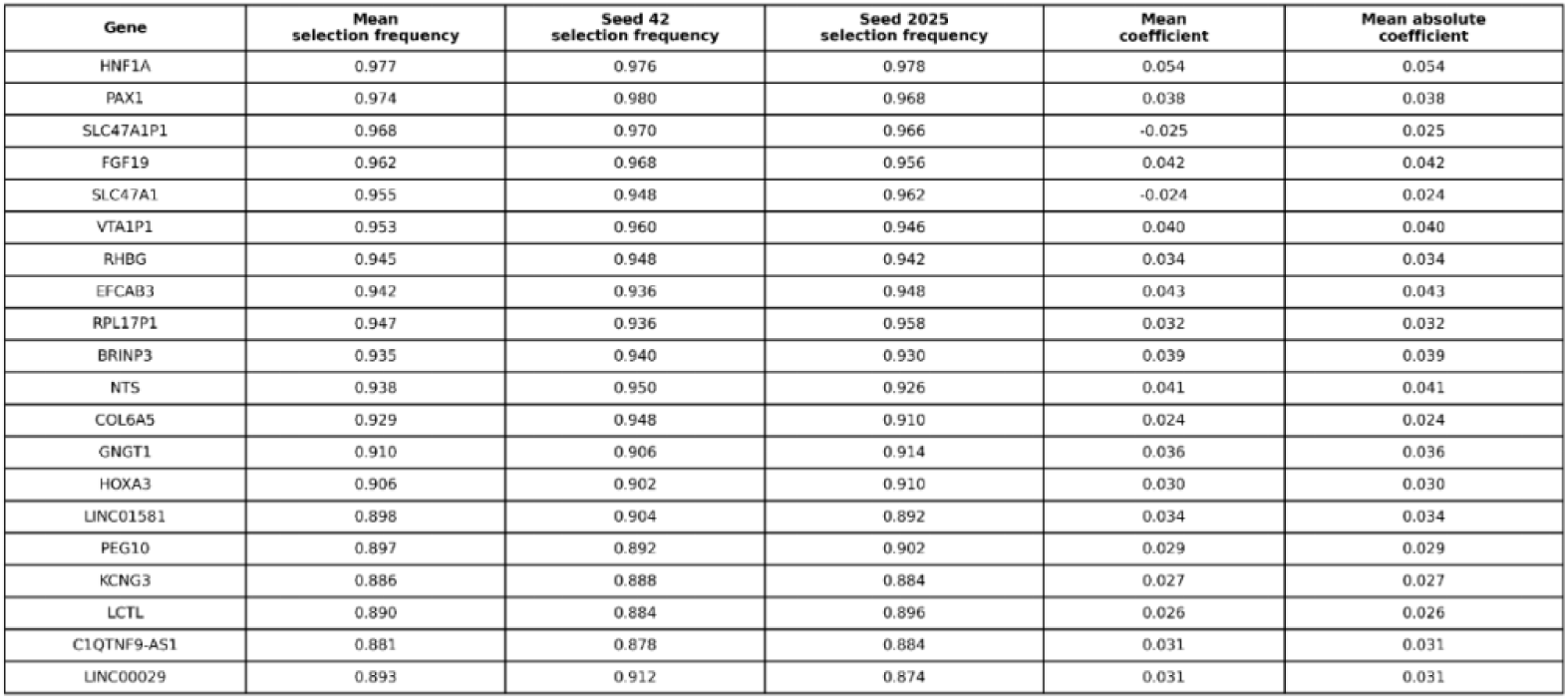
Core-stable genes (top 20) The top 20 core-stable genes ranked by mean selection frequency integrated across the two completed seed schedules are presented. Seed-specific selection frequencies and the integrated coefficient summary are provided together to numerically anchor the stability-based interpretation.

**Table 4.**
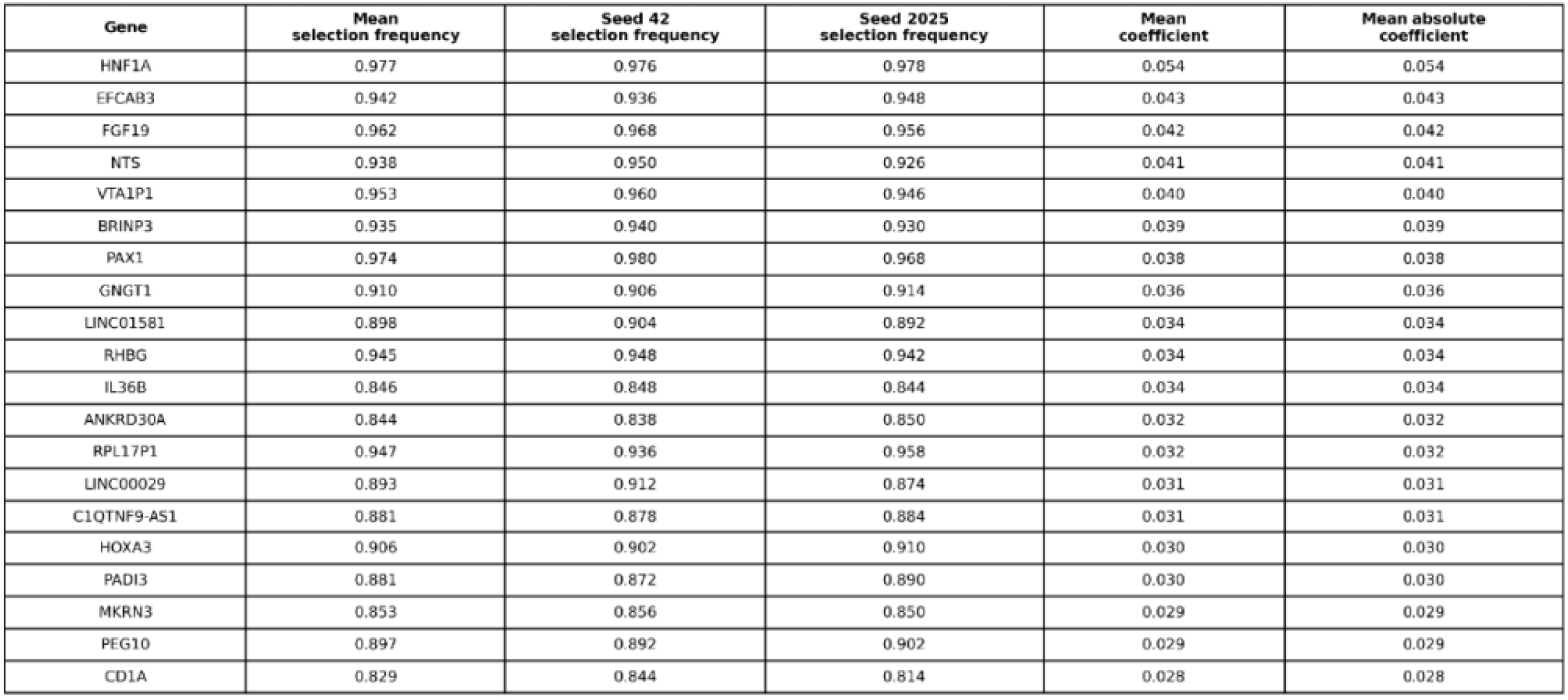
High-impact genes (top 20) The top 20 genes ranked by mean absolute coefficient magnitude in the integrated two-seed summary are presented. This table summarizes the genes with the largest mean effect sizes in the fitted classifier, independent of repeated selection frequency.

**Table 5.**
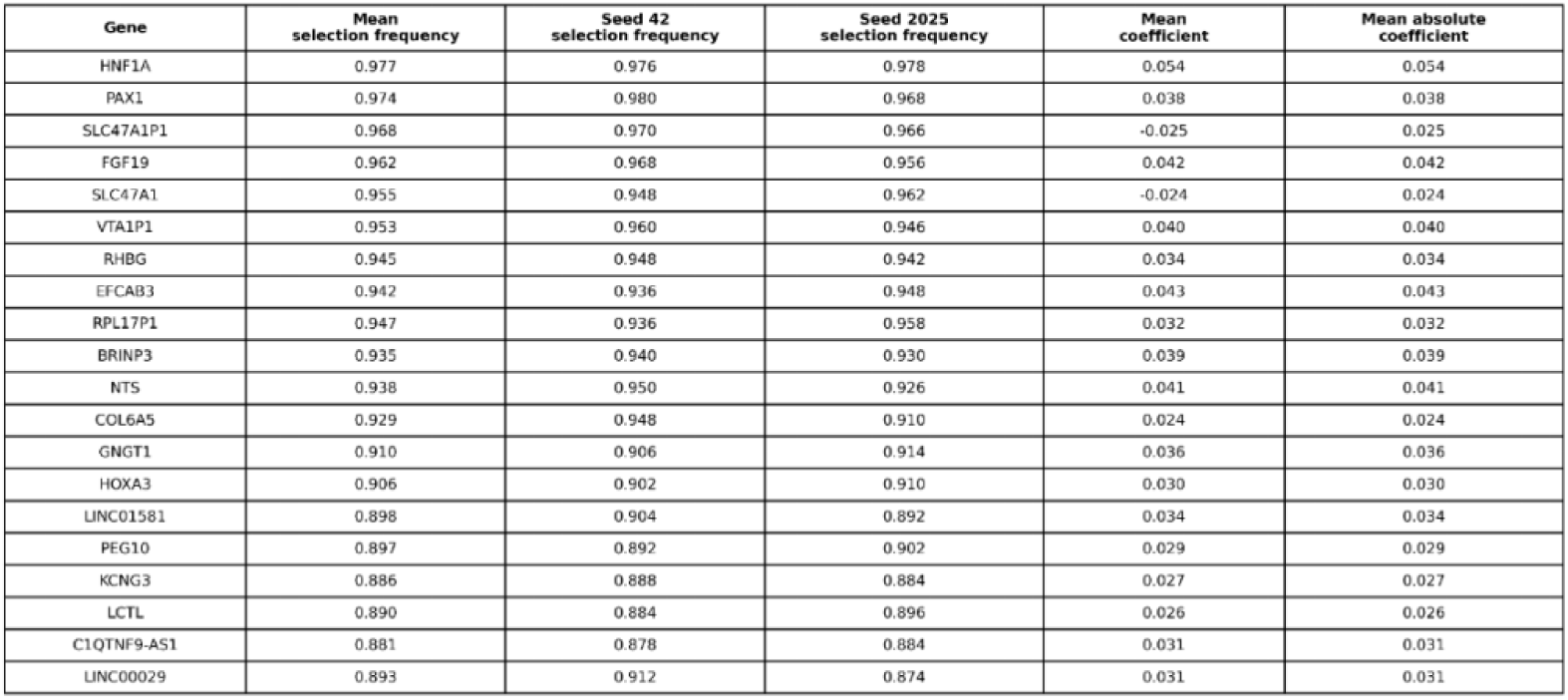
Overlap genes (top 20) The top genes belonging simultaneously to the core-stable set and the high-impact set are presented. This overlap set represents transcriptomic signals that are jointly supported by repeated selection frequency and large integrated coefficient magnitude.

**Table 6.**
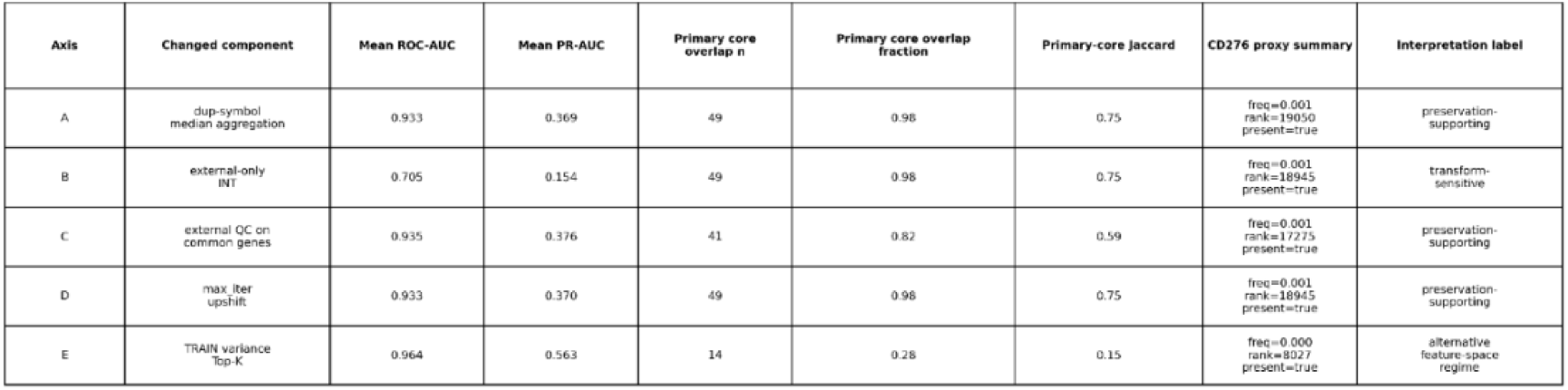
Robustness summary by axis This table summarizes, for each one-factor-at-a-time robustness axis, the changed component, mean external ROC-AUC, mean external PR-AUC, primary core overlap, primary-core Jaccard, CD276 proxy summary, and final interpretation label. It serves to numerically anchor the visual synthesis presented in Figure 4.

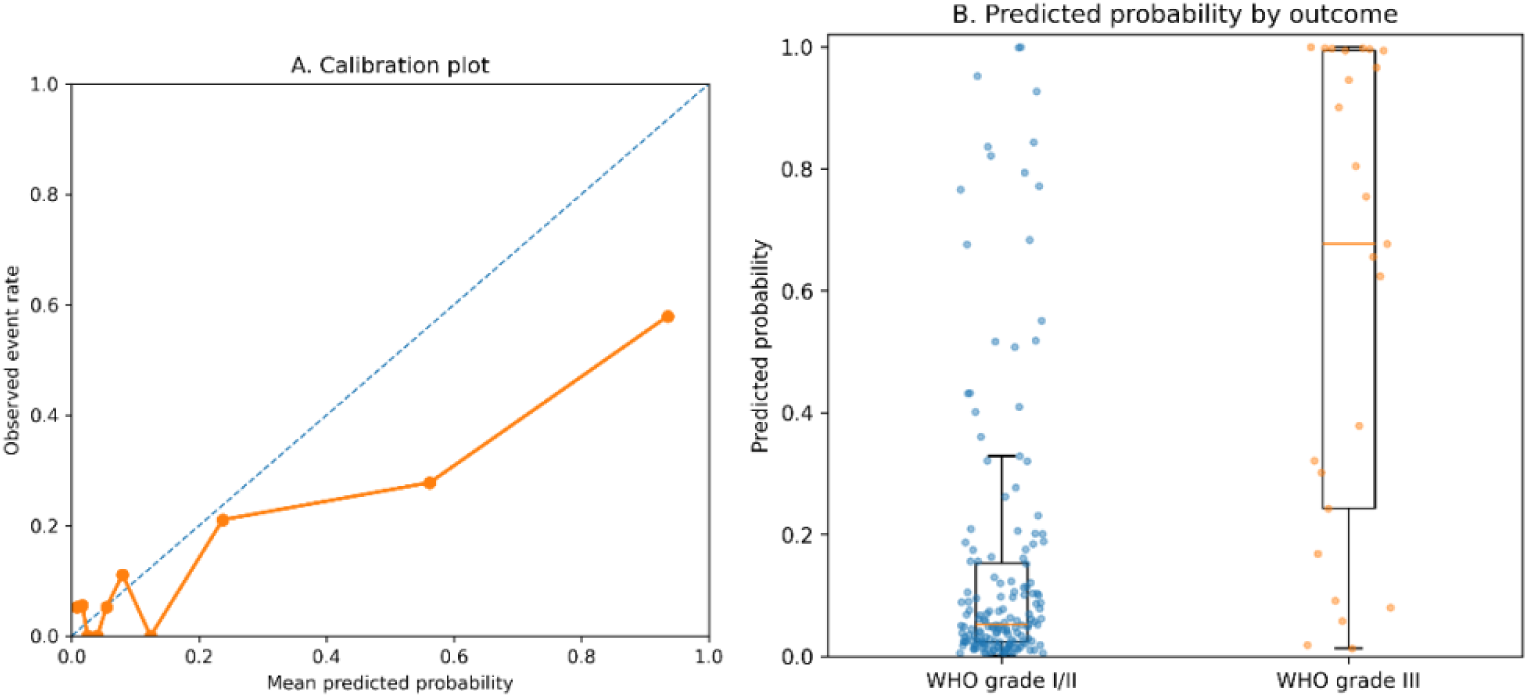

## Results-B

### External validation in a fixed common-gene space

We first restored the validated legacy analysis snapshot and re-established the aligned common-gene feature space shared by the training cohort and the external validation cohort. The restored state included 185 training samples with 25 positive cases and 160 external samples with 7 positive cases, all aligned to a fixed common transcriptomic axis of 31,582 genes. External validation was then reproduced in this restored feature space using a fixed L1-penalized logistic-regression pipeline.

In the external cohort, the classifier achieved ROC-AUC 0.928 and PR-AUC 0.335. At the default probability threshold used in the one-shot validation output, accuracy was 0.719 and balanced accuracy was 0.853, and the positive class showed complete recall of 7/7 at the cost of 45 false positives. These results suggest that the model maintained strong rank-based discrimination in the external cohort despite the low event prevalence. The bootstrap-based external validation summary was also consistent across two independent completed seed schedules. Across 500 bootstrap replicates, the mean ROC-AUC was 0.928 for seed 42 and 0.930 for seed 2025, and the mean PR-AUC was 0.352 and 0.354, respectively, supporting stable external discrimination at the performance level.

To evaluate whether this performance exceeded the level expected under a random label structure, we performed a train-label permutation null analysis evaluated on the external cohort. Under 1,000 deterministic permutations, the observed external ROC-AUC (0.928) and PR-AUC (0.335) both lay in the extreme upper tail of the corresponding null distributions, whose means were approximately 0.561 and 0.085, respectively. Therefore, the external discrimination observed in the restored pipeline was not readily explained by chance label alignment alone.

### Calibration and train-only threshold selection

Although discrimination remained strong in the external cohort, the raw external probabilities were not well calibrated. When the external probabilities stored in the restored snapshot were used, the raw model showed a Brier score of 0.221, a recalibration intercept of -3.378, and a recalibration slope of 0.300, indicating substantial deviation from ideal calibration.

To avoid bias arising from calibration based on external data, we fitted a logistic recalibration model using only the repeated out-of-fold predictions from the training cohort. This train-only recalibration fully preserved external discrimination (ROC-AUC 0.928; PR-AUC 0.335) while substantially improving probability calibration. After recalibration, the external Brier score improved from 0.221 to 0.052, and ECE10 improved from 0.380 to 0.110. The intercept and slope of the train-only recalibration model were -1.848 and 0.359, respectively, suggesting that the overly confident raw probability scale had been compressed.

Operating thresholds were selected using only the calibrated training out-of-fold predictions. The prespecified train-only rules yielded three operating thresholds: 0.17 for best F1, 0.15 for best Youden’s J, and 0.11 for the sensitivity-oriented rule, together with the reference threshold of 0.50. When these thresholds were applied once to the externally calibrated probabilities, the 0.17 threshold yielded 6 true positives and 25 false positives, corresponding to sensitivity 0.857 and specificity 0.837. Both the 0.15 and 0.11 thresholds recovered all 7 external positive cases, but at the cost of 39 and 79 false positives, respectively, whereas the conventional 0.50 threshold showed higher specificity (0.974) but lower sensitivity (0.286). Overall, these results demonstrate a clear tradeoff between maintaining sensitivity and the burden of false positives when thresholds are transferred from the training data to the external cohort. These results for external discrimination, calibration, and train-only threshold transfer are summarized in Figure 1.

### External decision-analytic utility

Next, we evaluated whether the train-only calibrated external probabilities showed clinically useful decision-making behavior. External decision curve analysis was performed over the threshold range of 0.01 to 0.50 together with patient-level bootstrap confidence intervals, and the displayed thresholds were fixed at 0.17, 0.15, 0.11, and 0.50, as selected by the train-only procedure.

Among these prespecified operating points, threshold 0.17 showed the most favorable external decision-analytic profile. At this threshold, the net benefit was 0.00550, whereas the Youden-optimal threshold of 0.15 was effectively near neutral (net benefit 0.00074), and both the sensitivity-oriented threshold of 0.11 and the conventional threshold of 0.50 were negative (−0.0173 and −0.0125). Thus, among the train-only candidate thresholds, only 0.17 showed a clearly positive external net benefit relative to treat-none.

Clinical-impact post-processing showed the same ranking. At threshold 0.17, the model corresponded to a net benefit of 0.55 per 100 patients and 76.95 net interventions avoided per 100 patients relative to treat-all. Threshold 0.15 reduced the number of interventions relative to treat-all but was close to decision neutrality, whereas thresholds 0.11 and 0.50, despite showing different false-positive burden patterns, were unfavorable from a net-benefit perspective. Taken together, these results suggest that external utility was not uniformly strong overall, but rather modest and threshold-dependent. The external decision-analytic profile is summarized in Figure 2.

### Stability-informed transcriptomic interpretation

We then shifted the focus from predictive performance to feature-level interpretability and used two completed stability-focused bootstrap runs performed on the validated common-gene axis. These runs used two independent seed schedules (42 and 2025), each consisting of 500 bootstrap replicates, while maintaining the same fixed modeling framework. External discrimination observed in the completed stability runs also remained similar to that of the main external-validation range, with mean ROC-AUC/PR-AUC of 0.931/0.370 for seed 42 and 0.935/0.372 for seed 2025.

Among the top-ranked genes in the core-stable set were recurrent genes such as HNF1A, PAX1, SLC47A1P1, FGF19, SLC47A1, VTA1P1, RHBG, EFCAB3, RPL17P1, and BRINP3, and the high-impact tier likewise emphasized HNF1A, EFCAB3, FGF19, NTS, VTA1P1, BRINP3, PAX1, GNGT1, LINC01581, and RHBG. The substantial overlap between the core-stable tier and the high-impact tier indicates that many of the most repeatedly selected genes were also those with the largest effect coefficients in the integrated summary across the two seeds.

In contrast, CD276 was not supported as a core stable classifier feature. In the integrated target-of-interest summary, the selection frequency of this gene was 0.002 for seed 42 and 0.000 for seed 2025, and it remained only in the target-of-interest tier rather than in the core-stable or high-impact groups. Thus, within this stability framework, CD276 is more appropriately interpreted as a biology-motivated follow-up target rather than as a dominant stable driver of the fitted classifier. The overall structure and key results of the stability-informed transcriptomic interpretation are summarized in Figure 3.

### Stability-informed enrichment analysis

To summarize the biological themes associated with the stability-informed gene sets, we performed enrichment analysis using a fixed common-gene universe of 31,582 genes. No pathways satisfied FDR < 0.05 in either the primary core-stable ORA or the secondary overlap-set ORA. Nevertheless, themes that repeatedly appeared among the top results across databases were observed. In the core-stable set, major GO and Reactome terms included amino-acid transport and SLC-mediated transmembrane transport, and top Hallmark terms included programs related to the G2-M checkpoint. The overlap-set analysis also showed similar qualitative patterns, including neuron-differentiation and neuropeptide-signaling themes, as well as transport-related Reactome categories.

Nominally significant enrichment after multiple-testing correction was observed only in the exploratory top-100 high-impact ORA, in which three GO Biological Process terms satisfied FDR < 0.05. In contrast, no Reactome or Hallmark categories in this exploratory set met the prespecified significance threshold. Likewise, the ranked stability-informed enrichment analysis performed across the full universe of 31,582 genes did not yield any FDR-significant results in GO, Reactome, or Hallmark. These results suggest that although certain biological themes appeared repeatedly, pathway-level signal remained generally limited under the fixed non-directional enrichment framework, supporting cautious interpretation.

### One-factor-at-a-time robustness analyses

Finally, we evaluated whether the primary interpretation was maintained across five prespecified one-factor-at-a-time robustness axes: duplicate-symbol aggregation (Axis A), external input transformation (Axis B), common-gene QC restriction (Axis C), convergence setting (Axis D), and feature prefiltering (Axis E). These robustness analyses were explicitly defined not as alternative primary analyses, but as stress tests of the fixed primary interpretation.

Three axes—A, C, and D—most strongly supported preservation of the primary biological interpretation. Axis A retained 49 of the 50 primary core genes (98.0%), Axis C retained 41 (82.0%), and Axis D retained 49 (98.0%). These axes also maintained similar mean external discrimination across seeds, with mean ROC-AUC/PR-AUC of 0.933/0.369 for Axis A, 0.935/0.376 for Axis C, and 0.933/0.370 for Axis D. From an integrative perspective, the primary core set showed Jaccard overlap of 0.754 with both Axis A and Axis D, and 0.587 with Axis C, suggesting that the primary anchor structure was substantially preserved.

Axis B occupied a distinct position. This axis retained 49 primary core genes (98.0%), thereby preserving anchor-gene structure at the gene level, but predictive discrimination decreased markedly under the altered external representation, with mean ROC-AUC 0.705 and mean PR-AUC 0.154. Thus, rather than suggesting overall biological instability, Axis B showed that predictive performance may be sensitive to external input representation even when anchor-gene structure remains broadly similar.

Axis E showed yet another pattern distinct from all other axes. This axis achieved the strongest discrimination among the robustness analyses, with mean ROC-AUC 0.964 and mean PR-AUC 0.563, but retained only 14 of the 50 primary core genes (28.0%). Its Jaccard overlap with the primary core set was also only 0.149. This pattern was closer to a different feature-space solution than to confirmatory preservation, although it was advantageous in terms of performance. Thus, the robustness results were not homogeneous. Some axes mainly supported preservation of the primary interpretation, Axis B revealed transform sensitivity, and Axis E exposed a qualitatively different high-performing feature regime.

Across these robustness analyses, CD276 did not emerge as a dominant robust core feature. The refined robustness framework positioned Axes A/C/D as preservation-supporting, Axis B as transform-sensitive, and Axis E as an alternative feature-space regime; within this structure, CD276 remained outside the dominant core top-ranked features, and its proxy signal was weakest in Axis E. The cross-axis robustness synthesis is summarized in Figure 4.

**Figure 4.**
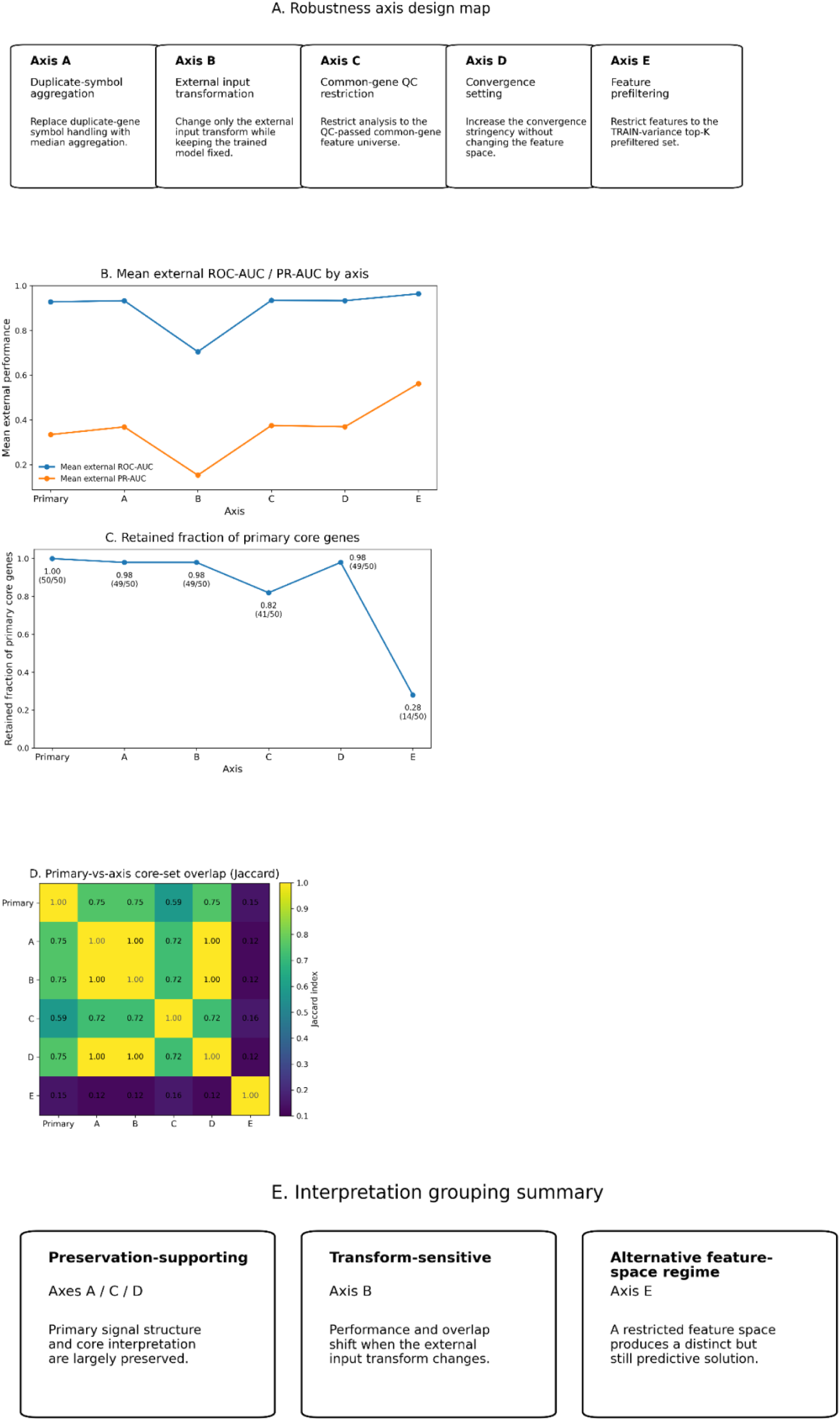
Cross-axis robustness synthesis. (A) Design map of the five one-factor-at-a-time robustness axes: duplicate-symbol aggregation (Axis A), external input transformation (Axis B), common-gene QC restriction (Axis C), convergence setting (Axis D), and feature prefiltering (Axis E). (B) Mean external ROC-AUC and PR-AUC by axis. (C) Retained fraction of primary core genes by axis. (D) Heatmap of Jaccard overlap between the primary core set and the axis-specific core sets. (E) Interpretation grouping summary, positioning Axes A/C/D as preservation-supporting, Axis B as transform-sensitive, and Axis E as an alternative feature-space regime. These robustness analyses were designed not as alternative primary analyses, but as stress tests of the primary interpretation.

## Discussion

This study sought to more rigorously clarify how CD276 should be interpreted by evaluating its position in meningioma transcriptomic data stepwise at the levels of the single-gene signal, multigene predictive modeling, external validation, and stability/robustness-based interpretation. In the internal training cohort, CD276 expression showed a significant association with WHO grade, and this directional pattern was maintained not only on the original TPM scale but also after log1p transformation and winsorization. However, the internal discrimination of the CD276 single-gene baseline classifier was limited, and recovery of grade III cases was also low. In contrast, the transcriptome-wide multigene model showed higher ROC-AUC, PR-AUC, accuracy, and balanced accuracy, and the CD276-forced 5,001-feature branch also maintained superior internal discrimination relative to the CD276-only baseline. These results suggest that although CD276 may be a gene of interest associated with meningioma grade biology, the main basis of predictive performance lies not in CD276 alone but in a broader multigene transcriptomic structure. In other words, the central conclusion of this study is that CD276 should be interpreted not as a strong standalone predictor, but within a broader transcriptomic program.

This interpretation did not rely solely on the simple comparison between single-gene and multigene performance. In the initial reduced feature space, CD276 was not included in the top 5,000 filtered gene set, and even in the forcibly included 5,001-feature branch, it remained at a low position in the permutation-importance ranking. Furthermore, in paired ablation repeated cross-validation comparing the CD276-including and CD276-excluding branches on the same splits, delta ROC-AUC, delta PR-AUC, delta balanced accuracy, and delta accuracy were all effectively close to zero. This more directly supports the view that CD276 is not a dominant feature that materially increases branch-level discrimination of the classifier. Therefore, the most appropriate way to interpret CD276 in this study is not as a standalone classifier or a core stable feature, but as a biology-motivated target-of-interest connected to the broader transcriptomic organization of meningioma.

An important point is that these internal results did not depend only on a specific split or a single setting. In the prediction-level bootstrap confidence intervals, the multigene branches consistently maintained a superior performance profile relative to the CD276-only baseline, and in the label permutation tests for the canonical full-transcriptome model and the 5,001-feature branch, the observed discrimination exceeded that expected under a random label structure. In addition, in the regularization-strength scan based on repeated stratified cross-validation, both multigene branches maintained generally stable performance ranges. These results support the conclusion that the internal findings of this study cannot be explained merely by chance splits or the choice of a particular C value, and that the limited contribution of CD276 and the superiority of multigene structure reflect a more structural phenomenon. In other words, the key signal observed in this study was not that “CD276 was meaningless,” but that “transcriptome-level organization beyond the CD276 single-gene signal was the substantive basis of predictive performance.”

The external validation results complemented this interpretation in an important way. Under a fixed common-gene axis and a fixed L1-penalized logistic-regression pipeline, the external cohort maintained a high ROC-AUC, and in the permutation-based external null analysis, the observed external discrimination also exceeded that expected under a random train-label structure. This suggests that the transcriptomic signal learned internally can substantially maintain rank-based discrimination in external data as well. In particular, although the CD276 single-gene signal was limited internally, the multigene classifier maintained strong discrimination externally, which is consistent with the interpretation that the predictive performance observed in this study was supported by a broader transcriptomic signal spanning the common-gene space rather than by any single specific gene. This is one of the most important integrative messages spanning both the internal and external results.

However, this study did not equate strong discrimination with the reliability of probability interpretation. In the internal stage, the OOF predicted probabilities of the initial transcriptome-wide multigene model showed an overconfident pattern, and in the external stage as well, the raw external probabilities were not well calibrated. Even though ROC-AUC and PR-AUC were maintained in the external cohort, the Brier score, recalibration intercept, and recalibration slope showed that the probability scale required separate correction. This became even clearer after train-only recalibration, as recalibration improved probability calibration while preserving discrimination. Accordingly, this study shows that although this model family may be sufficiently capable of ranking relative risk, calibration should be considered an essential post-processing step when interpreting the predicted probabilities themselves or applying them to threshold-based decision-making. Given that internal overconfidence and the need for external recalibration conveyed the same directional message, calibration may be regarded not as a secondary analysis in this study, but as a central element of model interpretation.

In this regard, an important methodological strength of this study is that both calibration and threshold selection were performed using a train-only strategy. To avoid optimistic bias arising from tuning based on external data, the recalibration model was fitted using only repeated out-of-fold predictions from the training cohort, and the operating thresholds were likewise derived solely from the calibrated training OOF probabilities. The recalibration and thresholds obtained in this way were then applied once to the external cohort. This design maximally preserves the independence of external validation while also allowing evaluation of practically usable threshold transfer and probability correction. Therefore, the external validation results of this study do not simply report high discrimination; they also have strong methodological credibility in that calibration and operating-point behavior were examined while minimizing leakage.

The results of decision curve analysis and threshold transfer showed that the practical utility of this model was threshold-dependent. When thresholds selected from train-only calibrated OOF predictions were applied to the externally recalibrated probabilities, not all thresholds showed the same utility, and a clear tradeoff existed between maintaining sensitivity and the burden of false positives. In other words, a high ROC-AUC did not automatically guarantee consistent clinical utility. Only some thresholds showed clearly positive external net benefit relative to treat-none, whereas other thresholds could be unfavorable from a decision-analytic perspective even when discrimination was maintained. These results emphasize that evaluation of a transcriptomic classifier cannot rely on ROC-AUC or PR-AUC alone, but should also present probability calibration, threshold transferability, and decision-analytic utility together. By reporting not only the strengths of the model but also the fact that utility was modest and threshold-sensitive, this study provides a more realistic interpretive framework.

From the perspective of feature-level interpretation, this study intentionally treated predictive performance and biological interpretation separately. In the stability-focused bootstrap analysis, a substantial overlap was observed between core-stable genes and high-impact genes, but CD276 was not supported as a dominant stable core feature and remained in the target-of-interest tier. This was also directionally consistent with the internal ablation results. In other words, CD276 is closer to a biology-motivated follow-up marker worth tracking beyond the classifier structure, rather than to the most stable and repeatedly selected core axis of the fitted classifier. This conservative positioning is also consistent with the interpretive principle of this study, which avoids equating predictive feature importance with biological relevance.

The enrichment analysis results also made this interpretation more cautious. Although some themes repeatedly appeared among the top results in the core-stable gene set and the overlap set, pathway-level signal was generally limited under a strict multiple-testing correction threshold. Some GO terms were significant in the exploratory high-impact ORA, but broad FDR-significant pathway enrichment was not observed in the overall framework, including ranked stability-informed enrichment. This suggests that feature-level stability or effect-size ranking does not directly translate into strong pathway-level significance. Therefore, for the biological interpretation of this study, it is more appropriate to remain with the conservative conclusion that although recurrent qualitative themes exist, their pathway-level statistical support is limited, rather than making strong claims that a specific pathway has been clearly confirmed.

The robustness analysis extended the interpretation of this study from simple repetition to structural validation. The five one-factor-at-a-time robustness axes did not all repeat the same message. Some axes strongly supported preservation of the primary interpretation, others revealed sensitivity to external input representation, and still others showed higher predictive performance while being closer to a qualitatively different feature-space solution. In other words, robustness did not mean that “the results were completely identical regardless of what was changed,” but rather showed that under some analytic perturbations the primary interpretation was preserved, under others sensitivity to representation was revealed, and in some cases an alternative but high-performing feature regime emerged. This asymmetry suggests that robustness should be used not as a simple confirmation of reproducibility, but as a tool for structurally understanding the boundaries and defensibility of the primary findings. From this perspective, this study considers it most appropriate to interpret Axes A/C/D as preservation-supporting, Axis B as transform-sensitive, and Axis E as an alternative feature-space regime.

This study has several limitations. First, because both the internal and external cohorts were based on public GEO data, the effects of sample size, event prevalence, metadata completeness, platform differences, and preprocessing heterogeneity cannot be completely excluded. Second, the interpretation of CD276 was based on its feature-level position, stability, coefficient behavior, and branch-level contribution within the transcriptomic classifier, and was not directly confirmed by protein-level expression or functional experiments. Therefore, although it is possible to position CD276 as a biology-motivated target-of-interest, extending this interpretation to the level of a therapeutic target or a direct mechanistic driver goes beyond the scope of the present study. Third, because pathway-level enrichment signal was limited, it is not appropriate to equate feature-level interpretation with pathway-level mechanism. Fourth, although external recalibration and threshold transfer were performed in a train-only manner to reduce leakage, additional validation in larger independent cohorts and prospective settings is needed before discussing actual clinical applicability.

In summary, this study shows that although CD276 is a gene of interest associated with WHO grade biology in meningioma transcriptomic classification, it is difficult to interpret it as a standalone predictor or a dominant stable classifier feature. When the internal and external results are considered together, the substantive basis of predictive performance lies not in CD276 alone but in a broader multigene transcriptomic structure, and probability output should be interpreted conservatively together with downstream processing including calibration and threshold selection. At the same time, stability and robustness analyses did not support CD276 as the strongest stable core driver, but instead positioned it in the target-of-interest tier within a broader transcriptomic organization. Therefore, this study proposes that CD276 is most appropriately understood not as a direct single-gene classifier, but as a biology-motivated follow-up marker worth tracking within the wider transcriptomic landscape of meningioma. Future studies should further evaluate how this signal may contribute to actual risk stratification and downstream biological investigation through larger independent cohorts, protein- and spatial-level validation, and prospective calibration and decision-utility assessment.

## Author Contributions

Hyojae Lee: Conceptualization, data curation, formal analysis, methodology, software, visualization, writing—original draft.

Hyeoneui Kim: Supervision, writing—review and editing, correspondence.

## Data Availability Statement

All datasets used in this study are publicly available from the Gene Expression Omnibus (GEO) under accession numbers GSE183653 and GSE136661.

## Code Availability Statement

The analysis code underlying this study is available from the corresponding author upon reasonable request.

## Ethics Statement

This study used only publicly available, de-identified datasets and did not involve new recruitment of human participants or direct interaction with patients. Therefore, institutional review board approval and informed consent were not required for this study.

## AI-Assisted Writing Declaration

The authors used generative artificial intelligence tools to assist with language editing, English translation, code generation, and manuscript drafting. The authors reviewed, revised, and accept full responsibility for the final content of the manuscript.

## Data Availability

All datasets used in this study are publicly available from the NCBI Gene Expression Omnibus (GEO) under accession numbers GSE183653 and GSE136661. All derived results necessary to interpret the findings are contained in the manuscript.

https://www.ncbi.nlm.nih.gov/geo/query/acc.cgi?acc=GSE183653

https://www.ncbi.nlm.nih.gov/geo/query/acc.cgi?acc=GSE136661

